# Artificial Intelligence in Biomedicine: Systematic Review

**DOI:** 10.1101/2023.07.23.23292672

**Authors:** Irene S. Gabashvili

## Abstract

Artificial Intelligence (AI) is a rapidly progressing technology with its applications expanding exponentially over the past decade. While initial breakthroughs predominantly focused on deep learning and computer vision, recent advancements have facilitated a shift towards natural language processing and beyond. This includes generative models, like ChatGPT, capable of understanding the ‘grammar’ of software code, analog signals, and molecular structures.

This research undertakes a comprehensive examination of AI trends within the biomedical domain, including the impact of ChatGPT. We explore scientific literature, clinical trials, and FDA-approval data, utilizing a thematic synthesis approach and bibliometric mapping of keywords to examine numerous subsets from over a hundred thousand unique records found in prominent public repositories up to mid-July 2023.

Our analysis reveals a higher prevalence of general health-related publications compared to more specialized papers using or evaluating ChatGPT. However, the growth in specialized papers suggests a convergence with the trend observed for other AI tools. Our findings also imply a greater prevalence of publications using ChatGPT across multiple medical specialties compared to other AI tools, indicating its rising influence in complex fields requiring interdisciplinary collaboration.

Leading topics in AI literature include radiology, ethics, drug discovery, COVID-19, robotics, brain research, stroke, and laparoscopy, indicating a shift from laboratory to emergency medicine and deep-learning-based image processing. Publications involving ChatGPT predominantly address current themes such as COVID-19, practical applications, interdisciplinary collaboration, and risk mitigation.

Radiology retains dominance across all stages of biomedical R&D, spanning preprints, peer-reviewed papers, clinical trials, patents, and FDA approvals. Meanwhile, surgery-focused papers appear more frequently within ChatGPT preprints and case reports. Traditionally less represented areas, such as Pediatrics, Otolaryngology, and Internal Medicine, are starting to realize the benefits of ChatGPT, hinting at its potential to spark innovation within new medical sectors.

AI application in geriatrics is notably underrepresented in publications. However, ongoing clinical trials are already exploring the use of ChatGPT for managing age-related conditions.

The higher frequency of general health-related publications compared to specialized papers employing or evaluating ChatGPT showcases its broad applicability across multiple fields. AI, particularly ChatGPT, possesses significant potential to reshape the future of medicine. With millions of papers published annually across various disciplines, efficiently navigating the information deluge to pinpoint valuable studies has become increasingly challenging. Consequently, AI methods, gaining in popularity, are poised to redefine the future of scientific publishing and its educational reach.

Despite challenges like quality of training data and ethical concerns, prevalent in preceding AI tools, the wider applicability of ChatGPT across diverse fields is manifest.

This review employed the PRISMA tool and numerous overlapping data sources to minimize bias risks.

## Introduction

### Rationale

The term Artificial Intelligence (AI) refers to systems capable at performing tasks that have traditionally required human intelligence –learning, problem-solving, and decision-making. AI’s transformative application has been widespread in theoretical medicine, affecting areas without direct patient contact such as drug discovery and - omics-based technology, pharmacology, and pharmacogenomics.

Despite AI’s significant potential to enhance and, in some cases, replace certain physician functions, its extensive integration into clinical medicine has faced obstacles. The sheer volume of scholarly articles and associated data generated in this field is overwhelming, and although there has been a marked increase in published research, valuable resources are being wasted due to ineffective utilization and the restricted impact of this research. Besides, many published scientific papers lack the maturity and direct applicability required by the healthcare industry, leading to a wealth of unstructured data and algorithms remaining underutilized [1].

In this paper, we analyze publications on AI in biomedicine, and emphasize the significant role of ChatGPT, an advanced language model, in enhancing communication and knowledge exchange within the field.

### Objectives

Our study primarily aims to evaluate existing literature related to ChatGPT against the backdrop of preceding AI tools. To inform a better understanding of the paradigm shift ushered in by ChatGPT, we focused on adoption trends and potential impacts across various biomedical fields. This is achieved through a systematic review and bibliometric analysis of primary literature.

Another objective involves uncovering patterns and potential avenues to analyze vast amounts of data efficiently, aiming to reduce waste (shown to be as high as 90% of published research) and enhancing the translation of research outcomes into practical applications. We also aim to investigate using ChatGPT for different stages of the review and synthesis process, with the goal of expediting the analysis of large volumes of scientific literature.

## Methods

### Study Design

This study follows the PRISMA guidelines [2]. The protocol is registered in PROSPERO [3], an international prospective register of systematic reviews at the National Institute for Health Research and the Center for Reviews and Dissemination (CRD) at the University of York (<u>CRD42023417336</u>).

The literature search spanned multiple databases, including Medrxiv, EuropePMC and PubMed. These platforms index research extensively in biomedicine and provide public accessibility. Building on our prior study that evaluated ChatGPT literature from December 2022 to April 2023 [4], we extended our search to encompass documents published up until July 15, 2023, guided by the search strategy previously delineated. To pinpoint subsets of documents pertaining to specific medical specialties, we adapted the queries by integrating relevant keywords, field of research and mesh terms.

### Research Questions

To examine the role and implications of AI tools, including ChatGPT, in the biomedical field, we aimed to address the following Research Questions (RQs):

RQ1: What applications of AI, including ChatGPT, exist in the biomedical sector, and how are these applications evolving?

RQ2: How do unique features differentiating ChatGPT from its AI predecessors contribute to a paradigm shift?

RQ3: What are the prevailing trends in AI application across the biomedical, clinical, and healthcare sectors?

RQ4: In which medical applications have AI tools, including ChatGPT, been trialed and evaluated?

RQ5: What are the perceived strengths, limitations, and main concerns associated with ChatGPT’s application in healthcare, especially in comparison to other AI tools and depending on the specific field of application?

RQ6: What significant research gaps exist or require further exploration in the existing literature concerning AI and ChatGPT usage in healthcare?

RQ7: Given the extensive volume of existing AI literature, what strategies and methodologies can effectively filter, categorize, and evaluate these documents in a manageable and meaningful way?

### Literature Search Strategy

To undertake a comprehensive review of literature on ChatGPT, we systematically identified and assessed all existing reviews in this domain. We also expanded our review scope to reviews of artificial intelligence in the biomedical and healthcare sectors. To address the significant volume of publications and ensure a thorough analysis, we implemented a method derived from the Capture-Recapture Technique. This involved selecting several overlapping subsets of unique records from multiple databases and subjecting them to repeated analyses, allowing us to navigate the vast expanse of literature with greater inclusivity.

The search strategy, developed across multiple publicly accessible databases and repositories, covered Medrxiv, EuropePMC, Pubmed, and Dimensions AI, filtered by 42 Health Sciences OR 32 Biomedical and Clinical Sciences Fields of Research, ANZSRC 2020 [5]. Additional sources included the Database of Artificial Intelligence and Machine Learning (AI/ML)-Enabled Medical Devices [6], patents [7], clinical trials registries, and protocols for systematic reviews database. We also incorporated specialized case report journals like Cureus, ensuring a broad sweep of literature types and annotations. The search strategy was adjusted for each database, applying the appropriate controlled vocabulary as required (for example, using mesh terms “family practice”[MESH] OR “primary health care”[MESH] in pubmed instead of “family medicine” or using label “Respiratory medicine” in medrXiv instead of Pulmonology)

To maintain consistency with our previous study [3], the same databases - Medrxiv, Biorxiv, EuropePMC, Pubmed, Dimensions AI, ClinicalTrials.gov, PROSPERO, Semantic Scholar, and Google Scholar - were screened using previously described search strategies.

Final search was conducted on July 15, 2023.

### Inclusion and Exclusion Criteria

To be eligible for inclusion, the studies needed to be research papers that reviewed either ChatGPT or other artificial intelligence topics in the biomedical domain. We specifically looked for systematic reviews of reviews that focused on artificial intelligence techniques in healthcare and had their protocols registered in the PROSPERO database. Due to ChatGPT’s recent introduction, we did not exclude reviews of this tool that did not meet all of the PRISMA and AMSTAR criteria. However, we only selected reviews that provided a clear explanation of their publication selection process. We were more stringent with artificial intelligence reviews considering only reviews with published protocols in PROSPERO database.

### Study Selection

The selection process of reviews was carried out in a similar way to reviews of individual studies. Additionally, subsets of primary publications were selected from each database and compared to each other. The PRISMA flow diagram was used to detail the literature screening and selection process.

The initial screening involved evaluating titles and abstracts, followed by a comprehensive review of potentially relevant studies in full text. To ensure relevance and avoid duplication, the titles were carefully verified and any identical titles across different versions of the same article were identified and eliminated, particularly preprint versions. In cases of repeat publication, priority was given to the most recent and comprehensive data.

### Data Extraction & Synthesis

In order to effectively explore extensive collections of scientific literature, we concentrated on identifying prevalent themes, evolving trends, types of publications, and research areas.

We gathered data on MeSH terms, author keywords, tags allocated by the journal or the publication database, key subjects, Fields of Research, Publication Date, and Publication Type. We also collected the specifics of the literature search, sources of data, and the outcomes of research or systematic review. The main outcomes were advantages and disadvantages of utilizing Artificial Intelligence tools, such as ChatGPT, within the healthcare sector.

Data collection from each report was conducted by the human author, while ChatGPT independently extracted data and assigned keywords. Any inconsistencies were resolved through iterative reviews and improved prompt engineering, in order to reach a consensus. Claude and Bard were also used for independent labeling and keyword extraction tasks.

For a comprehensive understanding of the research landscape, we performed subset analyses focusing on specific kinds of articles like systematic reviews, case reports, and preprints. This analysis allowed us to pinpoint overrepresented domains within each subset, thereby highlighting potential research gaps, biases, or areas of excessive focus.

In assessing the quality of the research, we examined the criteria used for review selection and inclusion. Additionally, we conducted publication bias assessments, discussed heterogeneity test results, and compared the eligibility criteria, study characteristics, and primary outcomes of interest across the included reviews.

For keyword-based clustering, we utilized common words, labels, or topics to examine the content and categorizations within the publications. This strategy expedited the detection of pertinent clusters and patterns.

To combine key discoveries from the reviewed papers, we adopted a thematic synthesis approach. To analyze and visualize the organization, interactions and evolution of scientific knowledge, we used VOS Viewer software [8].

## Results

### Outcomes

The paper selection process pertaining to ChatGPT is illustrated in Figure 1 via a PRISMA flow diagram specifically designed for updated systematic reviews. This investigation included a total of 27 reviews focused on ChatGPT. Of these, 15 reviews were not exclusively targeted at the biomedical sector, but rather addressed areas such as general education (6 reviews), societal impacts (7 reviews), business process supply chains (1 review), and information security (1 review). Notably, all of these papers either explicitly addressed the impacts of ChatGPT in medicine. Of the remaining 12 reviews, the majority (7 reviews, [9–15]) explored the general healthcare field, with the rest being related to medical education (1 review, [16]) or specialized medical areas such as dentistry [17,18], obstetrics and gynecology [19], dermatology [20], or public health [17] (4 reviews).

**Figure 1.**
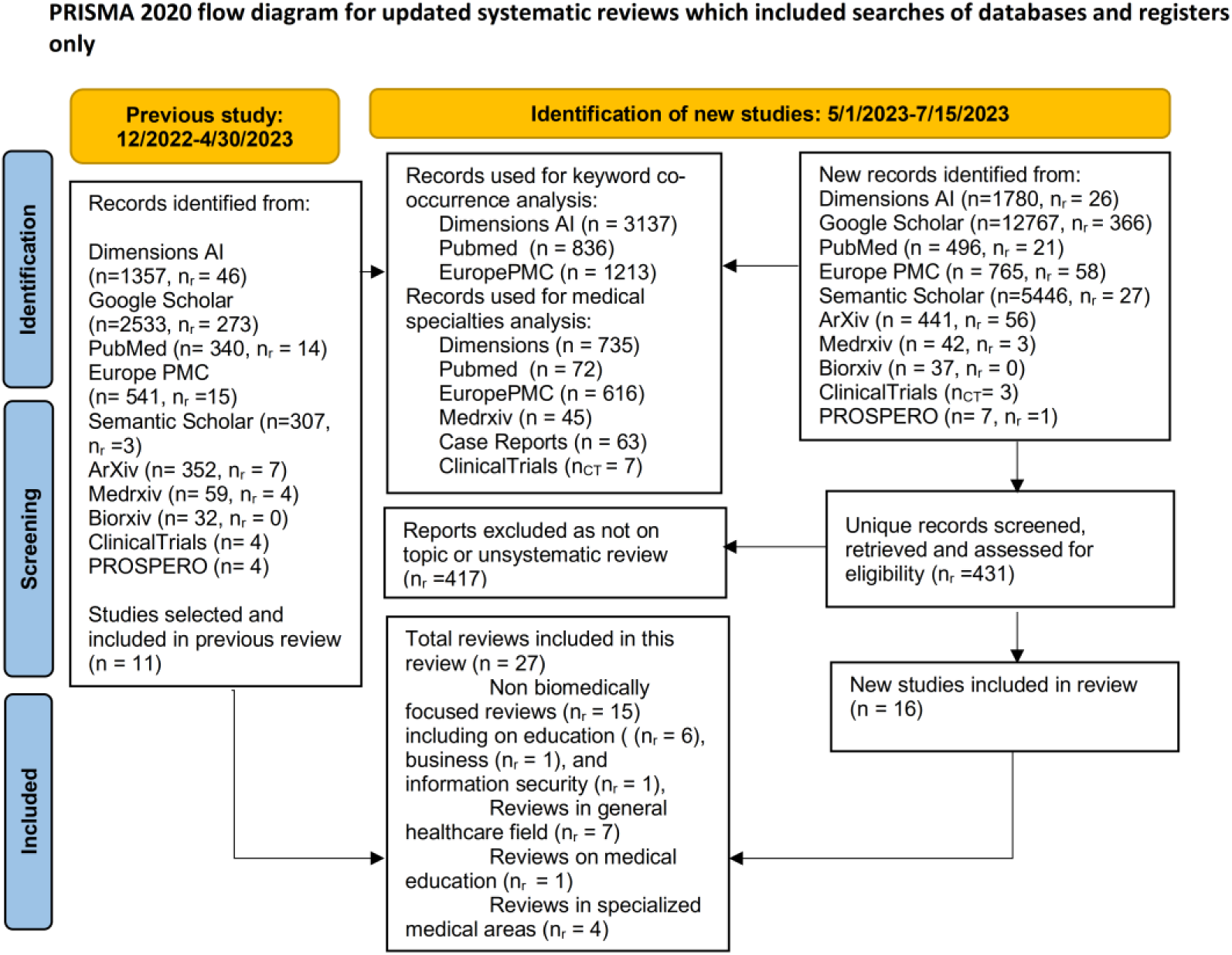
Selection of ChatGPT publications

Figure 2 visualizes the PRISMA flow diagram for new systematic reviews on artificial intelligence, showcasing the selection process for the articles. This review encompassed a total of 77 AI-focused systematic reviews. In the figure, the notation ‘n’ refers to the number of original papers identified in various databases covering all years up until July 15, 2023. The ‘nt’ symbol represents the articles incorporating the term ‘artificial intelligence’ in their titles, ‘nr’ signifies the number of reviews, ‘2023’ in subscript refers to papers published in 2023, and ‘bh’ in subscript denotes the number of papers specifically filtered from the biomedical and health fields. In this AI review, multiple subsets of articles were selected for VOSviewer analysis and visualization, with their counts ranging from sever al thousand to tens of thousands.

**Figure 2.**
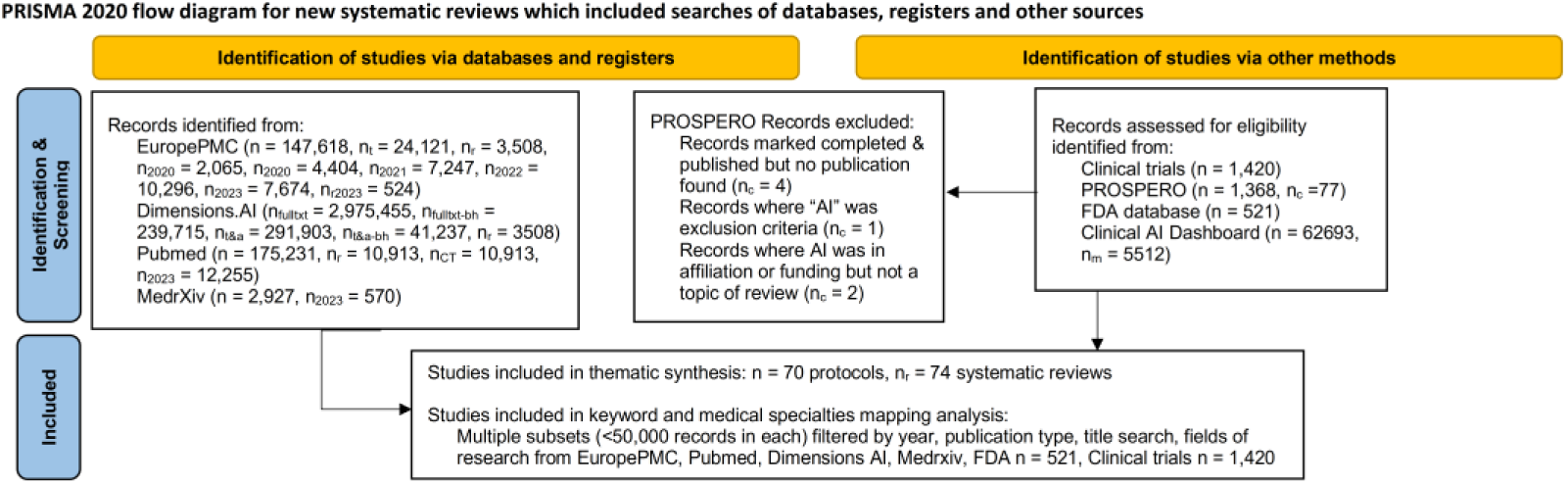
AI publications

The thematic synthesis analysis incorporated a total of 74 systematic reviews on AI applications in healthcare and 27 reviews on ChatGPT applications, including 12 reviews concerning the biomedical and healthcare fields. Our analysis found multiple themes related to medical specialties, identified limitations, and potential future directions. However, for the purpose of this review, we have chosen to concentrate on the predominant theme - the outcome. We classified the outcome of systematic reviews into the following categories:

- Reliable: Indicating effective use of the AI tool, with sensitivity and specificity within the range acceptable for clinical use. This category might include studies concluded that the use is still limited - reliable in certain applications within a given medical specialty, but not others.
- Promising: While it may still have issues, the tool was deemed to have considerable potential.
- Unreliable: insufficient predictive ability of AI algorithms
- Inconclusive: Findings did not definitively place the tool in any of the above categories.

As per our findings, all reviews on ChatGPT could be categorized as “Promising”. The outcomes for AI reviews were as follows: 60% were deemed “Reliable” and either already in use or ready for utilization, 28% were considered “Promising” (marginally effective, potentially effective, or not yet effective but showing promise), 3% were classified as “Unreliable”, 5% were “Inconclusive”, and 4% were either underutilized or not utilized at all despite its potential.

### Medical Specialties

Figure 3 presents a heatmap displaying the distribution of medical specialties across preprints (sourced from MedrXiv and Dimensions.AI, labeled as Biomedical and Clinical Sciences and Health Sciences), CASE REPORTS (predominantly from Cureus), systematic reviews with PROSPERO protocols, clinical trials, patents, and papers from PubMed and EuropePMC. Notably, the table indicates a significant overrepresentation of radiology and oncology across all subsets. Emergency medicine is adequately represented only among case reports. Unlike other AI tools, ChatGPT has made inroads into additional fields - it is significantly represented in internal medicine, pediatrics, and otolaryngology. Gerontology is underrepresented in all subsets, with the exception of clinical trials involving the use of ChatGPT.

**Figure 3.**
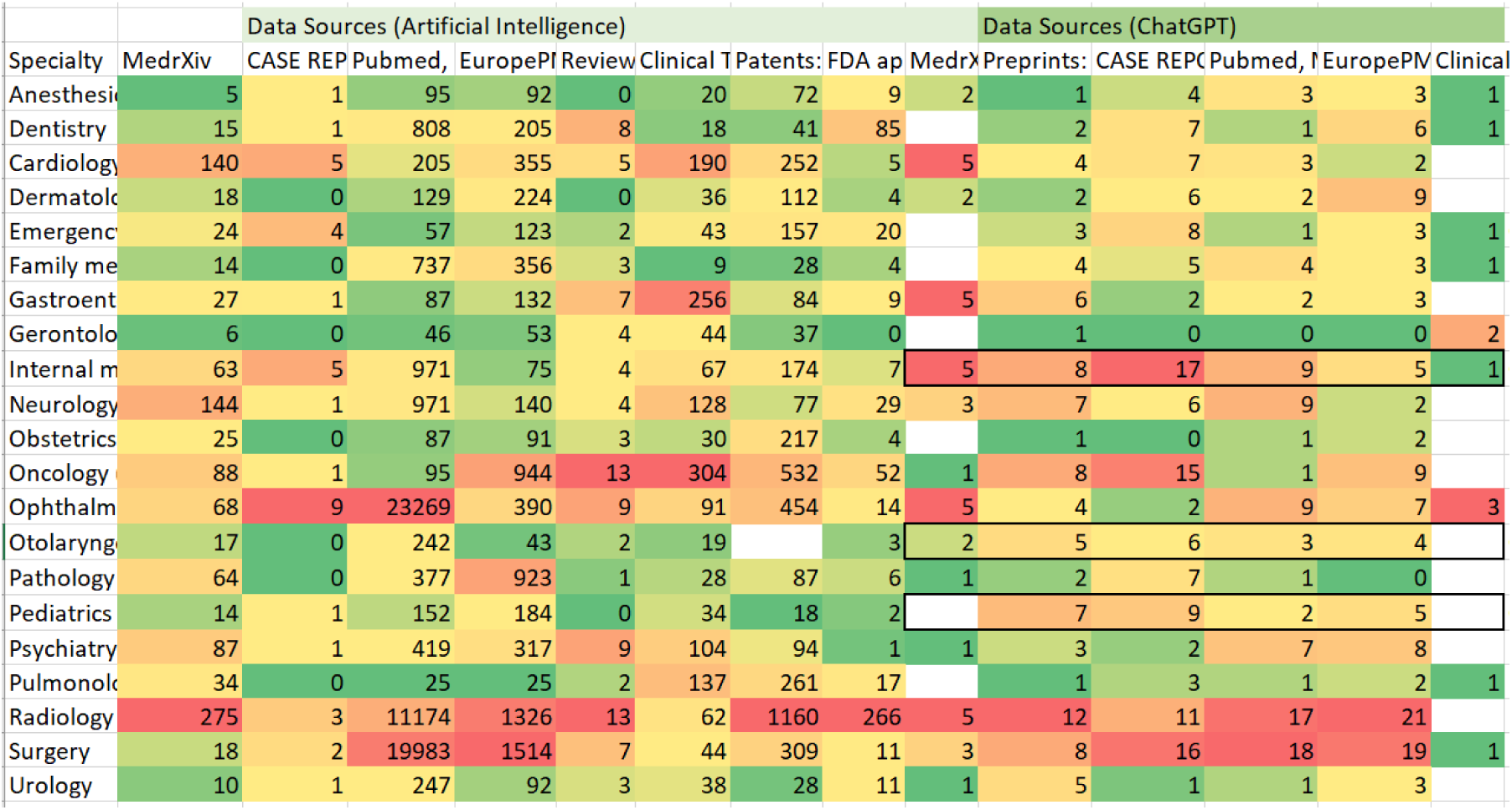
Heatmap of AI and ChatGPT use by medical specialties

**Figure 4.**
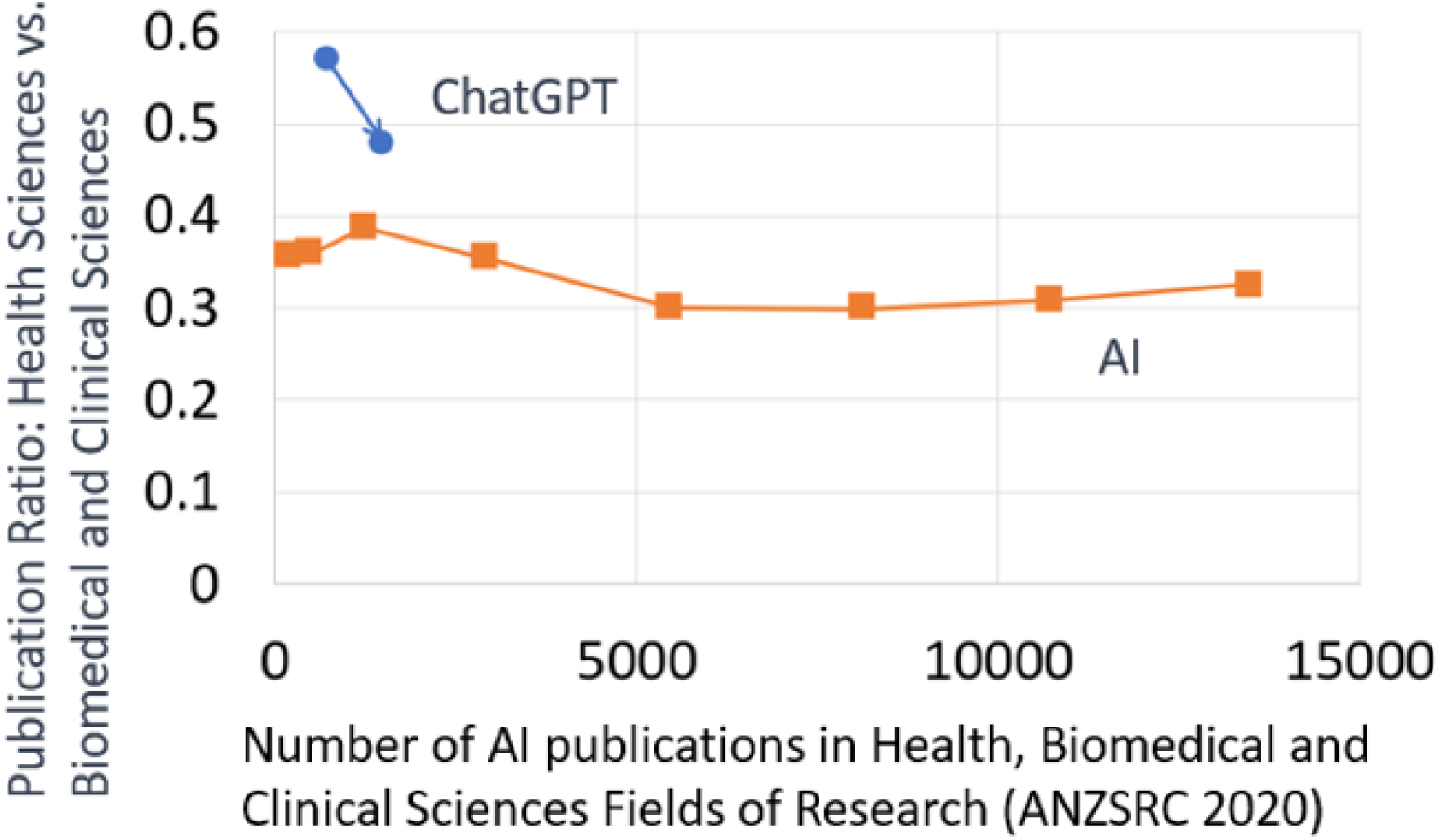
Ratio of AI and ChatGPT publications in general healthcare field vs biomedical and clinical fields

At the point of our previous review [4], the proportion of ChatGPT publications within the health sciences category (according to the 42 Health Sciences Fields of Research delineated by Dimensions.AI’s ANZSRC 2020 [5]) surpassed that within the Biomedical and Clinical Sciences category (which consists of 32 Fields of Research). The ratio of Health to Biomedical and Clinical publications stood at 0.57.

However, two and a half months later, as the total count of ChatGPT publications within these categories surpassed the 1,000 mark, the ratio dipped below 0.5. This brought it closer to the ratio seen for AI publications. This shift, indicating a growing number of specialized papers relative to more general health publications, signifies the ongoing evolution of ChatGPT’s role within the health domain.

### Analytical themes

The analysis of keyword co-occurrences revealed major thematic clusters in AI publications over the years, denoting the evolving focus of research in the field. In EuropePMC AI papers, the dominant themes in 2014 through 2016 were reproducibility of results and computational biology, followed by medical informatics and biomedical research, and later, brain, robotics, telemedicine, big data and synthetic biology. New themes in 2017 were delivery of healthcare and neurological models. 2018 saw the rise of publications about using AI in wearable electronic devices, genomics (especially for breast and lung cancers), precision medicine and personalized medicine (with the latter term less popular than the former). AI continued to be explored in the delivery of health care and decision making. In 2019 radiology became more prominent, and delivery of healthcare was often mentioned along with clinical decision support. The year 2020 witnessed a diversification of themes with an emphasis on colonoscopy, ophthalmology, pediatrics, precision medicine, radiotherapy, and the brain. In 2021, the focus, again, shifted towards the reproducibility of results, COVID-19, drug discovery, breast cancer, mammography, wearable devices, and coronary artery disease. Computational biology re-emerged as a key theme of AI-guided drug discovery and deeper understanding of disease. In 2022, the recurring themes included COVID-19, drug discovery, lung and colon cancers, mental health, digital health, AI ethics, and diabetic retinopathy. By 2023, the major clusters revolved around precision medicine, radiomics, delivery of healthcare, COVID-19, reproducibility of results, ethics, and ChatGPT.

**Figure 5.**
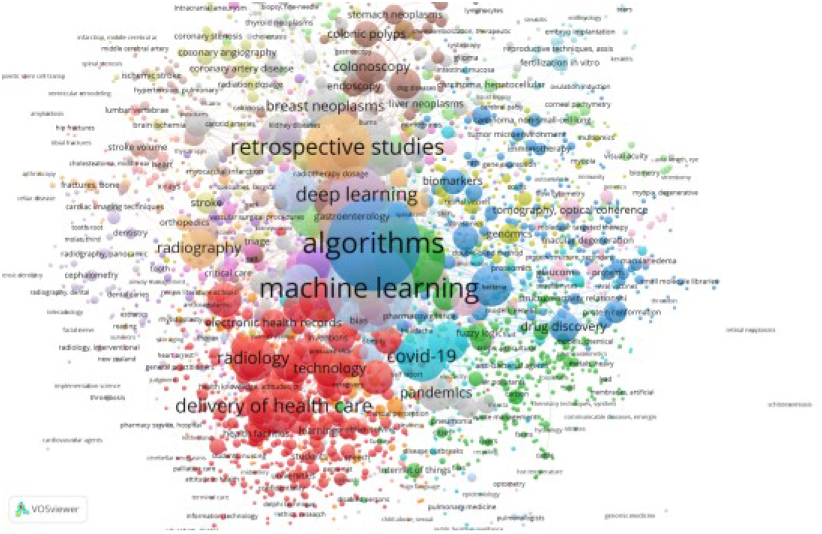
Keyword co-occurrences for papers with AI in their title

Analysis of clinical trial papers published in PubMed highlighted themes such as robotics, exercise therapy, stroke rehabilitation, postoperative complications, laparoscopy, coronary artery disease, facial expressions, and urinary incontinence. Furthermore, PubMed reviews clustered around the brain, laparoscopy, computational biology, and stroke rehabilitation, with additional emphasis on cardiovascular diseases, cancers, and surgical procedures.

The temporal analysis of AI literature demonstrated the evolving popularity of certain keywords and the increasing penetration of AI into different medical specialties. Earlier focus on artificial intelligence, artificial neural networks, and big data transitioned towards machine learning, deep learning, and ultimately, large language models (LLMs) such as ChatGPT. Ethics emerged as a theme preceding the advent of LLMs.

In PubMed Case Reports mentioning AI tools, initial focus was on computer-assisted surgery, computer-assisted diagnosis tools and computer simulation, with prevalent medical conditions being diabetes and cardiovascular diseases. Later, these reports started to reflect themes such as reproducibility of results, phenotype, and traumatic brain injury. Since 2020, COVID-19 has heavily dominated these reports. More recent themes include robot-assisted surgery, cardiac ultrasound, emergency medicine, automatic seizure detection, telemedicine, pregnancy, genetic mutations, and protein diagnostics.

FDA approvals over time show the earliest endorsements for AI were in critical care medicine, pathology and laboratory medicine, followed by radiology, neurology, physical therapy, cardiology, gastroenterology, pulmonology, and urology. The latest approvals revolve around advanced deep-learning-based image and signal processing, as well as MRI.

**Figure 6.**
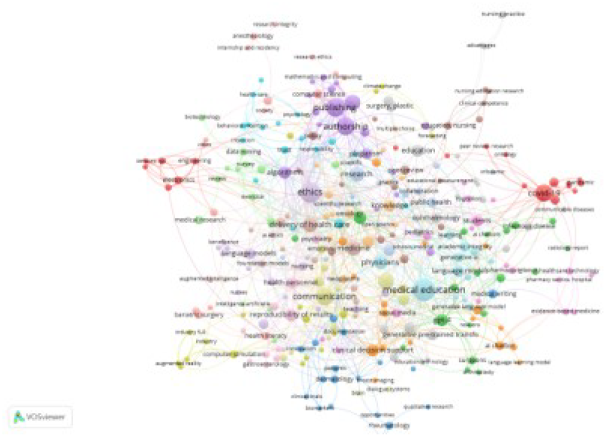
Keyword co-occurrences in ChatGPT publications

Analyzing ChatGPT themes with Dimensions.AI biomedical and health subsets revealed major keywords such as future, bias, medical education, case, radiologist, trust, test, examination, complexity, decision support, plagiarism, short-learning, and prompt engineering. In EuropePMC, major themes included medical education, COVID-19, ethics in association with publishing and authorship, clinical decision support, IOT and sensors, plastic surgery, rheumatology, occupational health, radiology, delivery of healthcare, plastic surgery, physicians, and a smaller cluster for computational biology. PubMed analysis revealed dominant themes around medical education and ethics, publishing and authorship, and reproducibility of results, with smaller clusters focusing on nursing education, emergency medicine, plastic surgery and bariatric surgery.

## Discussion

### Principal Findings

The key findings of this systematic review underscore ChatGPT’s substantial potential in transforming the medical landscape. Despite the existing constraints cand limitations, this AI tool has demonstrated successful application in areas such as authoring case reports from electronic health records [20], conducting systematic reviews [14, 20, 21], generating search queries, analyzing abstracts and full texts, and effectively classifying and labeling information. It also showed potential in fostering awareness and combating misinformation.

Healthcare involves a plethora of intricate tasks that have traditionally been challenging to automate. Generative AI models such as ChatGPT mark a pivotal shift in handling this complexity. These AI solutions are gradually permeating into more complex and specialized areas of biomedicine, helping to streamline processes and improve efficiency.

Historically, the integration of AI tools in healthcare necessitated considerable investment in training healthcare professionals. This precondition often instigated resistance towards adopting such technologies, especially if the benefits were not immediately apparent. However, ChatGPT seems to be dismantling these obstacles. It facilitates user-friendly interaction and delivers tangible benefits, rendering it more accessible and attractive to healthcare professionals. Therefore, the current findings provide robust evidence for the sustained development and application of ChatGPT and similar AI tools within the biomedical and healthcare industries.

The higher prevalence of general health-related publications relative to more specialized papers that utilize or evaluate ChatGPT is indicative of its broader applicability across multiple fields.

### COVID-19

The COVID-19 pandemic, a prominent theme throughout the reviewed literature, has provided an unprecedented context for the practical application and evaluation of AI tools, including ChatGPT. These tools have been employed for more than just image diagnostics and writing tasks, expanding to include research question identification, filtering of scientific literature, screening health records, responding to healthcare queries, providing contextual information, and aiding data analysis.

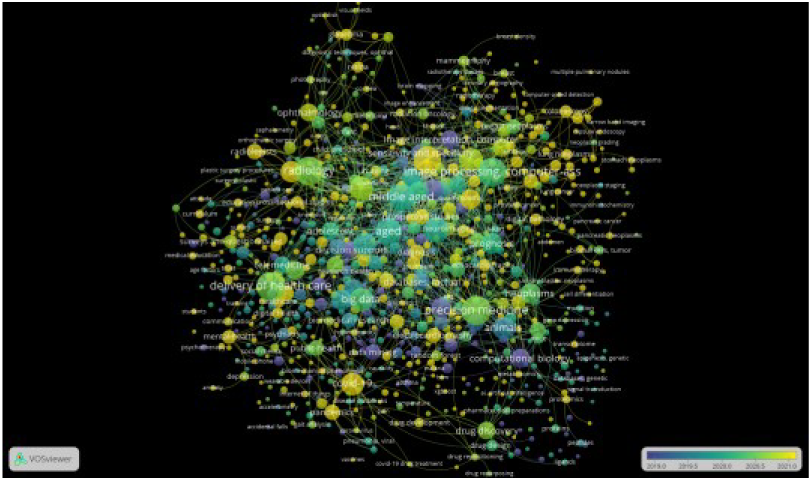

An analysis of 70 PROSPERO protocols was conducted as part of this systematic review, with five being centered on COVID-19. Three studies demonstrated the effectiveness of AI algorithms in tasks such as COVID-19 detection from high-resolution CT images [22], patient triaging based on medical imaging [23], and conducting systematic reviews [24]. However, these studies also highlighted considerable limitations related to data heterogeneity and the limited quantity of available data for AI training, indicating a requirement for enhanced future research [23]. A fourth study [25] reinforced the need for high-quality datasets, reproducible studies with thorough documentation, and external validation to increase the likelihood of these models being integrated into future clinical trials.

Conversely, this same study [25] indicated that, despite extensive research efforts to develop machine learning models for COVID-19 diagnosis and prognosis, there are significant methodological shortcomings and biases present in the literature. This results in an overly optimistic representation of performance. A fifth study in the COVID-19 subset of reviews [26] suggested the potential effectiveness of AI in predicting mortality, screening, tracing, analyzing health data, and resource allocation in healthcare settings during pandemic.

In summary, the full effectiveness of AI tools has yet to be realized due to issues such as methodological flaws, prevalent biases in the literature, a shortage of high-quality datasets, and inadequate documentation for reproducibility and external validation [25].

The intersection of ChatGPT and COVID-19 is explored in over 100 papers, showcasing a range of potential applications. Despite the evident heterogeneity across these studies, a consistent potential is apparent. Particularly within community-based studies, unconstrained by large pharmaceutical funding or traditional academic boundaries [27], the use of AI tools like ChatGPT can offer considerable benefits.

This systematic review analyzed 12 systematic reviews focusing on the application of ChatGPT in biomedical, clinical, and health-related fields [9–20]. Notably, half of these reviews referenced COVID-19 in some capacity: two included COVID-19 in discussion, while another three indirectly examined ChatGPT papers related to the pandemic.

Review [15] noted that even though the growth of literature on ChatGPT is impressive, the rapid growth in literature pertaining to COVID-19 overshadowed it.

Review [18] found that ChatGPT could provide accurate responses to queries relating to a range of medical topics including medical examinations, systematic reviews, clinical reasoning, diagnostic imaging, liver diseases, and COVID-19 vaccination. Another review [10] further emphasized ChatGPT’s utility in analyzing population-level data on vaccine effectiveness, COVID-19 conspiracy theories, and compulsory vaccination.

A case report mentioned in review [20] detailed how ChatGPT was utilized to process information about a patient recently immunized with the COVID-19 vaccine. ChatGPT accurately assessed the patient’s skin condition, which was warm, dry, and free of rashes or erythema, and successfully ruled out dermatological and autoimmune causes for the reported symptoms of epigastric pain and heartburn.

### Case Reports

ChatGPT’s exceptional capabilities across a range of medical domains enable a streamlined process for both generating [28] and understanding medical reports [29].

It has been proposed that ChatGPT can yield coherent, comprehensive, and clinically pertinent medical reports with high levels of accuracy and consistency. The precision and reliability of the generated reports have been further validated through comparison with suggestions derived from an online doctor consultation system [28].

ChatGPT’s utility in drafting case reports could aid not only with the writing process, but also in the collection and dissemination of knowledge that would otherwise remain unreported, supporting community-based studies operating on limited budgets, and assisting healthcare professionals who often lack the time to draft case reports. Case reports, which play a crucial role in the discovery of novel findings, facilitating learning, and providing clinical guidance, often remain unpublished due to the challenges involved in effectively articulating observations. ChatGPT helps to address these challenges [20], aiding physicians in accurately communicating their findings and sharing valuable clinical insights.

In many patient cases, a multidisciplinary understanding spanning multiple medical specialties is critical. ChatGPT meets this need by enabling physicians to quickly understand topics with which they might be less familiar. While practitioners must still verify the information’s accuracy, ChatGPT significantly reduces the time required for this verification. For instance, in MRSA cases, pediatricians have found ChatGPT particularly useful when an infectious disease specialist is not part of their team [30]. Even though these case series were primarily labeled under categories like pediatrics and infectious disease, they still required expertise from infectious disease, radiology, orthopedics, cardiology, neurology, critical care medicine, and dermatology. Similarly, a complex case report assisted by ChatGPT [31] covered multiple disciplines, even though it was categorized under Internal Medicine, Radiology, and General Surgery by the publisher. It extended to Emergency Medicine, Infectious Disease, Radiology, Plastic Surgery, Oncology, and Genomic Medicine. ChatGPT demonstrated its capacity to take an electronic medical record written with multiple abbreviations, shortened forms of words, and incomplete ideas and sentences. It was able to rewrite the record in a way that expanded all of the shortened language into full, clear sentences and completed the unfinished thoughts to create a comprehensive and coherent document.

However, a notable challenge lies in the existing publishing model for medical case reports. Despite the numerous preprint servers available for medical publications, case reports are only accepted by journals that charge submission and publication fees. Even for less expensive options such as Cureus, these fees can pose a financial burden for some community-led initiatives.

The preference in medical literature for positive case outcomes adds another layer of complexity. This preference, likely driven by the fear of legal repercussions linked to negative case outcomes, results in a significant number of educationally beneficial cases going unreported and hence lost. Platforms like ResearchGate, which hosts a small fraction of such case reports (eg, [32]), and Open Science Framework (OSF), could potentially offer a better solution, fostering a more comprehensive and inclusive understanding of medical cases.

### Scientific Publishing

The fact that many educationally-valuable case reports fail to get published and are therefore lost to future clinicians is one major shortcoming of current medical publishing models. However, this is not the only problem with modern research literature practices.

Recent bibliometric analysis of biomedical literature showcased a growing prevalence of AI-assisted writing in peer-reviewed journals, even before the widespread use of ChatGPT [33], contributing to an exponential increase in publications. The accelerated rate of research publication over recent decades, though beneficial to overall scientific advancement, has also led to a glut of superficial articles lacking qualitative or quantitative evaluations [4.9].

ChatGPT’s review [19] found that papers describing a study carried out—typically more complex and evidence-based—have a lower impact factor than other publications (mean 6.25 ± 0 vs. 25.4 ± 21.6, p < .001).

The use of BERT-based analysis to assess the maturity level of research publications based on their abstracts revealed that levels of maturity are low and keep decreasing [34]. Out of 7062 AI-related healthcare publications from 2019–2021, only 385 were classified as mature. While 6.01 percent of publications in 2019 were mature, this figure rose to 7.7 percent in 2020 before falling to 1.81 percent in 2021. Radiology led with the highest number of mature model publications across all specialties in the past three years, followed by pathology in 2019, ophthalmology in 2020, and gastroenterology in 2021.

An evaluation through the Translational Evaluation of Healthcare AI (TEHAI) framework [35] revealed a consistent trend: studies often score highly for technical capability but underperform in areas vital for clinical translatability. Concerns regarding external model validation, safety, nonmaleficence, and service adoption generally received failing scores in most studies.

The current system incentivizes researchers to publish more, sometimes resorting to result fabrication. It also encourages publishers to accept lower-quality articles as authors bear the costs. The existence of paper mills and citation farms, which multiply publications and citations for a fee, combined with the promotional efforts of large institutions, highlight the dire need for an overhaul of scientific publishing practices.

The publishing industry currently acts as an intermediary, selecting papers for publication based on potentially biased peer reviews and favoring those who can afford to pay, which inherently introduces bias. Predatory journals, infamous for their absence of stringent peer review and editorial services, are not the only culprits; reputable journals also exhibit biases, favoring certain topics, authors, institutions, or regions.

Currently, the publishing industry serves as an intermediary, choosing papers for publication based on potentially prejudiced peer reviews and favoring those who can afford to pay. This model inherently introduces bias. Predatory journals, notorious for their lack of robust peer review and editorial services, are not the sole offenders; reputable journals also exhibit biases, favoring certain studies, authors, institutions, or regions.

Moreover, the rise of populism in scientific publishing is worrisome. A misplaced focus on a study’s potential popular appeal often results in less flashy but crucial work being sidelined. The presence of ‘paper mills’ and ‘citation farms’, which mass-produce and cite papers for profit, exacerbates this issue, compromising the integrity of academic publishing.

The situation is further complicated by the rapid increase in the volume of scientific publications. As millions of papers are published annually across numerous disciplines [34], sifting through the information overload to identify pertinent and valuable studies has become progressively difficult. Therefore, AI approaches, growing in popularity, represent the future of scientific publishing and its educational potential. These approaches can help ensure a comprehensive, representative selection of content and expedite the processing of relevant content.

### Limitations

The sheer volume of available literature on artificial intelligence within the realm of medicine presents a limitation. Although we utilized techniques to comprehensively analyze and navigate the vast array of publications, our review does not capture the entirety of the available literature. Our meta-review of AI in biomedicine was confined to the review of papers with protocols published in the PROSPERO database. While our review of reviews on ChatGPT is extensive and encompassing, the rapidly evolving nature of the field suggests that our findings may not fully encapsulate all studies concerning the application of ChatGPT in biomedicine and healthcare.

The accuracy and comprehensiveness of our findings are contingent on the quality and completeness of the original publications. The variability among the included reviews in terms of their focus, scope, methodology, and quality may undermine the applicability of our results to a wider context.

Another limitation is that this systematic review was conducted solely by a human author, which introduces the potential for subjective interpretations.

### Comparison with Prior Work

To our knowledge, this is the first and only systematic review of reviews that explores the applications and implementation of ChatGPT in biomedical, clinical, and health domains. It builds upon and extends our previous work on the impact of ChatGPT across various domains and industries [4]

While several reviews of reviews examining AI in the healthcare domain exist, they focus on specialized topics, including ethics, regulatory challenges, AI in orthopedics, the role of AI in the COVID-19 response, and pediatric oncology [36–40].

One meta-review [36] illuminated the ethical complexities linked with AI utilization in healthcare, probing into critical issues such as patient privacy, informed consent, transparency, fairness, data bias, inequity, and the “black box” phenomenon. The authors advocated for an enlargement and classification of the prevailing ethical framework. Echoing this sentiment, our review of ChatGPT also underscored the imperative for ethical innovation [4].

Another study [37] analyzed 16 reviews on the regulation of AI in Digital Radiology. This paper emphasized the need for a robust regulatory framework, highlighting the necessity for an international collaboration, heightened transparency and interdisciplinary bias-free methodologies.

Trend analysis of reviews analyzed in our paper, showed the growing emphasis on the same issue in ChatGPT publications.

Reviews [38] and [39] spotlighted AI applications in orthopedics and the COVID-19 pandemic respectively, with the latter underscoring the rising prominence of deep learning models and generative AI.

**Figure 7.**
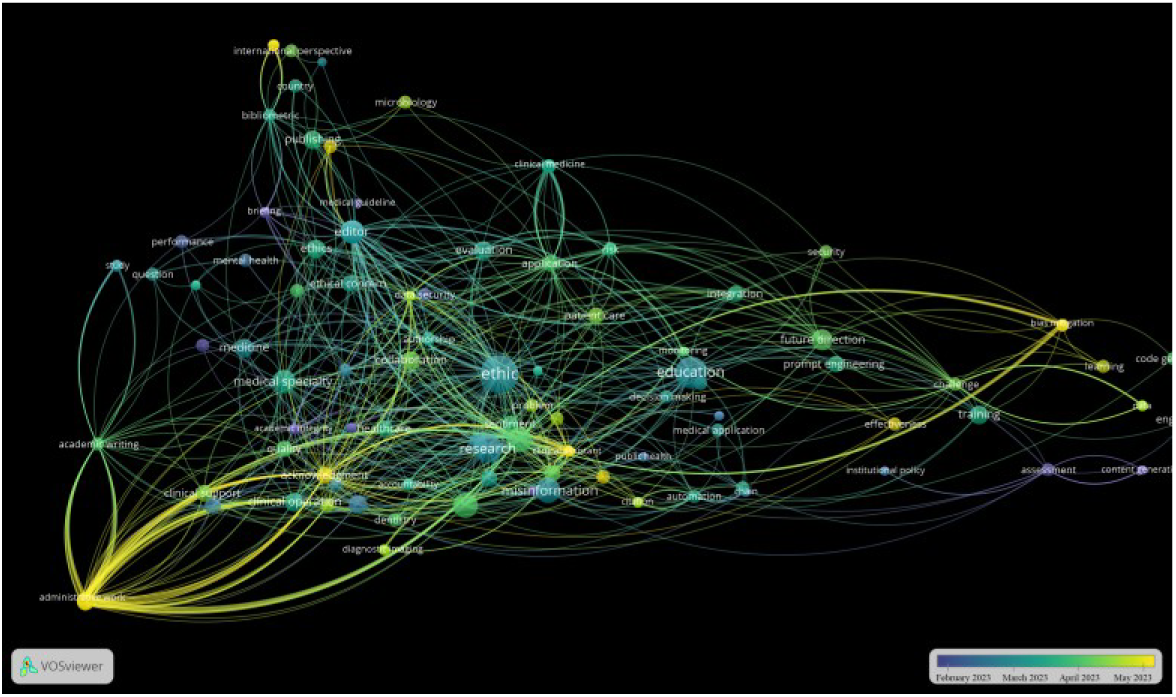
Keywords in ChatGPT reviews colored by the final date of literature searches

In pediatric oncology, a review [40] highlighted the embryonic stage of AI application but recognized its potential to address several unmet needs in the field.

Our review underscores the pioneering role of ChatGPT in fostering the adoption of AI tools within pediatrics and other areas that have been historically underrepresented in AI application.

### Conclusions

This systematic review of reviews highlights the rapidly expanding incorporation of AI technologies in biomedical, clinical, and health research fields. In particular, ChatGPT has acted as a catalyst, fostering significant strides especially within previously underrepresented areas.

Our review unveils a transformative shift in the publication landscape of AI within medicine, notably regarding ChatGPT. While initial studies and reviews were largely centered on broad health-related domains, the emerging abundance of specialized research signifies the progressive sophistication of ChatGPT within the healthcare arena. Emerging themes in ChatGPT research align with the prevailing trends seen in broader AI studies.

Predominant themes weaving through AI literature involve radiology, ethics, drug discovery, and the sustained influence of COVID-19. Meanwhile, newer exploratory topics encompass emergency medicine, robot-assisted surgery, personalized medicine, wearable devices, and -omics-based diagnostics.

Radiology maintains a robust influence across all phases of biomedical research and development, mirroring trends observed in general AI applications. However, the rising frequency of interdisciplinary-focused papers within ChatGPT preprints and case reports attests to its burgeoning relevance across diverse fields. The potential advantages of ChatGPT are becoming increasingly apparent within traditionally less represented disciplines such as Pediatrics, Otolaryngology, and Internal Medicine, underscoring the technology’s potential to ignite innovation across novel medical frontiers.

While challenges such as the quality of training data, trust in AI and ethical considerations are ubiquitous across all AI tools, including ChatGPT, all reviews acknowledge potential mitigation strategies and areas for refinement. Our study reiterates the pressing need to address ethical issues and to formulate robust regulatory frameworks.

In conclusion, AI, and more specifically, ChatGPT, hold promising potential for transforming the medical landscape. As we mitigate challenges and refine applications, we can look forward to groundbreaking advancements in patient care and medical research.

## Data Availability

All data produced in the present study are contained in the manuscript or available online.

https://osf.io/87u6q/

## Acknowledgements

The author thanks ChatGPT for assistance. The author also acknowledges gaining useful feedback from other AI systems: Bard, Bing, Claude, and SciSpace Copilot.

## Data Availability Statement

Data is available as open access at OSF: https://osf.io/87u6q/

## Conflicts of Interest

None declared.

### Abbreviations

AI: Artificial Intelligence
AMSTAR: Assessing the Methodological Quality of Systematic Reviews
DL: deep learning
FoR: Field of Research
GPT: generative pre-trained transformer
LLM: Large Language Model
MeSH: Medical Subject Headings, a controlled and hierarchically-organized vocabulary produced by the National Library of Medicine.
ML: machine learning
OSF: Open Science Framework, a platform for data sharing and collaboration
PRISMA: Preferred Reporting Items for Systematic Reviews and Meta-Analyses
VOSviewer: “Visualization of Similarities Viewer” - a software tool developed for constructing and visualizing scientific landscapes

